# Conus Medullaris Position in 9,808 Pediatric Lumbosacral MRI Examinations: A Large-Cohort Reference Distribution and the Normally Positioned Conus in Surgically Treated Tethered Cord

**DOI:** 10.64898/2026.06.06.26355031

**Authors:** Wenwen Tang, Yuan Dong, Jinliang Chen, Yue Yang, Hong Huang, Mengyan Yu, Jun Zhu, Gang Shen

**Author notes:** **Correspondence:** Gang Shen.

## Abstract

**Background:** Tethered cord syndrome (TCS) is classically associated with a low-lying conus medullaris, yet many surgically treated children have a normally positioned conus (“occult” TCS). Large-scale normative data on conus position in children, and the diagnostic value of quantitative conus assessment, are limited.

**Purpose:** To establish a large-cohort reference distribution for conus medullaris termination level in children, to quantify conus position in children surgically treated for presumed (occult) TCS, and to test whether automated conus segmentation and radiomics can distinguish TCS from normal.

**Materials and Methods:** In this retrospective single-center study, conus termination level was extracted from structured radiology reports of consecutive pediatric lumbosacral MRI examinations and encoded numerically (L1 = 1, L2 = 2, …). Children surgically treated for tethered cord were identified by linkage to an operative registry (name and date of birth) and restricted to preoperative examinations. A deep-learning model (nnU-Net) was trained for conus segmentation on axial T2-weighted images. IBSI-compliant radiomic features were extracted; reproducibility was assessed by intra- and inter-observer intraclass correlation (ICC). A case–control radiomics analysis used batch-only ComBat harmonization and cross-validated L1-penalized logistic regression; discrimination was compared with conus level by paired bootstrap.

**Results:** Among 9,808 examinations with a parseable conus level (98.5% of reports; parser validated against dual blinded annotation, 99.4% agreement, κ 0.946), the conus terminated in the L1 region in 85.7% and the L2 region in 14.3% of the reference cohort (postoperative examinations excluded, n = 9,655); a low-lying conus (≥L3) occurred in only 0.05% (5/9,655), and remained rare (0.14%, 14/9,808) including operated examinations (median L1; mean 1.13 ± 0.33). A slightly more cephalad position was seen with increasing age (negligible correlation). Among 475 preoperative children surgically treated for tethered cord, 99.6% had a normally positioned conus (≤L2) and only 0.4% were low-lying. Automated conus segmentation achieved a held-out Dice of 0.85. Conus radiomics likewise did not distinguish TCS from controls (equivalence-tested null; full segmentation/radiomics pipeline reported in the companion methodological paper).

**Conclusion:** In children, the conus medullaris terminates at L1–L2 in more than 99% of cases and is normally positioned in virtually all children surgically treated for TCS. Within the conus, neither position nor texture (radiomics) identifies tethered cord; whether the filum terminale carries a diagnostic signal was not tested here.

**Key Results:**

- In a reference cohort of 9,655 children (postoperative examinations excluded; conus level by a validated parser), the conus terminated in the L1 region (85.7%) or L2 region (14.3%); a low-lying conus (≥L3) occurred in only 0.05% (0.14% including operated examinations).
- Among 475 preoperative children surgically treated for tethered cord, 99.6% had a normally positioned conus, consistent with a radiologically occult presentation.
- Automated conus segmentation (Dice 0.85) was deployed cohort-wide; quantitative conus texture (radiomics) likewise did not discriminate tethered cord (full pipeline in the companion paper).

## Introduction

Tethered cord syndrome (TCS) is a clinico-radiologic disorder in which abnormal caudal fixation of the spinal cord produces progressive neurologic, urologic, and orthopedic dysfunction. The classic imaging hallmark is a low-lying conus medullaris, typically defined as a conus tip below the L2–L3 disc space, frequently accompanied by a thickened or fat-infiltrated filum terminale [1–3]. Because the conus normally ascends during fetal and early postnatal development, conus position has long served as a principal imaging criterion for the classic, low-lying form of the diagnosis, used alongside filum abnormalities rather than in isolation [4–8].

In practice, however, a substantial proportion of children who undergo surgical detethering have a normally positioned conus on preoperative MRI — a presentation often termed “occult” tethered cord. The existence and management of occult TCS remain debated, in part because the diagnosis rests on subtle filum abnormalities and clinical findings rather than on conus position [1–3]. Reliable, large-scale reference data on conus position in children are scarce; most published reference ranges derive from small cohorts or adult populations, limiting their applicability to pediatric practice [4–8].

Quantitative image analysis and radiomics have been proposed as a means of detecting tissue changes that are invisible to the eye, raising the possibility that texture analysis of the conus might identify tethered cord even when the conus appears normal [9,12]. Whether such signal exists has not been tested in a large, confound-controlled pediatric cohort.

In this study we leveraged a large single-center registry of pediatric lumbosacral MRI examinations to (a) establish a large-cohort reference distribution for conus termination level across childhood, (b) quantify conus position in children surgically treated for tethered cord identified through operative-registry linkage, and (c) evaluate whether automated deep-learning conus segmentation and IBSI-compliant radiomics can distinguish tethered cord from normal.

## Materials and Methods

Study design and ethics. This retrospective study was approved by the Ethics Committee of Women and Children’s Hospital of Ningbo University (approval no. NBFE-2026-KY-130; expedited review; approved 2 June 2026), which waived informed consent; examinations were performed between January 2021 and May 2026. Consecutive children (<16 years) who underwent lumbosacral MRI at a single tertiary pediatric center between January 2021 and May 2026 were eligible.

### Data sources

Three linked sources were used: (1) a radiology information-system export of structured reports (clinical indication, findings, and impression); (2) the institutional operative registry of spinal procedures, including patient name, date of birth, and operation date; and (3) the corresponding DICOM image archive.

### Conus termination level from reports

Conus termination level is routinely stated in our reports in standardized phrasing (e.g., “the conus medullaris terminates at the lower border of the first lumbar vertebra”). Levels were extracted with a rule-based parser and encoded on a continuous vertebral scale (T12 = 0, L1 = 1, …, with ±0.3 for upper/lower vertebral margins). An improved parser (resolving non-standard phrasing used for abnormal/low-lying and postoperative cases) succeeded in 9,808 of 9,956 examinations (98.5%) and was validated against independent dual blinded human annotation in a random sample of 361 examinations (exact agreement 99.4%; weighted κ 0.946; inter-rater κ 1.00; low-lying positive predictive value 100% after correcting one sacralization-related parsing error). A low-lying conus was defined as a tip at or below L3 (≥ 2.7). Postoperative examinations were excluded when deriving the reference distribution.

Surgically treated tethered cord (operative cohort). Patients in the operative registry with a tethered-cord– related procedure were linked to imaging by match on name and date of birth (±30 days), after de-duplicating the operative registry to unique patients. To avoid postoperative normalization, analyses of conus position were restricted to examinations performed before the operation date (preoperative subset, n = 475). Surgery was indicated on clinical grounds (symptoms together with filum findings); this operative cohort therefore represents children treated for presumed tethered cord and is not an independent, imaging-based reference standard. Procedures were predominantly filum sectioning/detethering, with a minority involving lipoma- or meningocele-related repair; analyses stratify primary (filum-type) versus lesion-associated tethering. Surgical indications were based on multidisciplinary clinical assessment, including progressive neurological, urologic, orthopedic, or cutaneous findings together with filum or dysraphic MRI features.

### Image processing and series selection

DICOM data were converted to NIfTI (dcm2niix). For segmentation and radiomics, an axial T2-weighted series covering the conus was selected automatically on the basis of plane orientation and geometric coverage.

### Automated conus segmentation

A 2-D nnU-Net (version 2) was trained to segment the conus medullaris on axial T2-weighted images using manually annotated cases, with iterative active-learning refinement. Segmentation accuracy was assessed on a held-out validation set (n = 152): Dice 0.85 (IQR 0.78–0.92), 95% Hausdorff distance 5.7 mm (median), average symmetric surface distance 0.98 mm (median), and conus-tip cranio-caudal localization within one slice thickness in 76% of cases. The trained model was then applied across the cohort and used to generate uniform segmentations for the case–control radiomics analysis, so that cases and controls were segmented identically.

### Radiomic features and reproducibility

IBSI-compliant features were extracted from the conus volume (MIRP; 1-mm isotropic resampling; fixed bin number, 32 bins). Reproducibility was assessed by intra- and inter-observer re-segmentation (30 examinations each); features with a two-way random, single-measure ICC ≥ 0.75 in both analyses were retained as stable (68 features).

### Harmonization, modeling, and statistics

To remove scanner/sequence effects without information leakage, ComBat harmonization was applied using sequence as the only batch variable (no outcome covariate). Discrimination of TCS from controls used L1-penalized logistic regression within repeated stratified five-fold cross-validation; the conus level and a combined (radiomics + level) model were evaluated identically. AUCs were compared with a paired bootstrap (2,000 resamples). Analyses used Python (scikit-learn, neuroCombat). P < .05 was considered significant. Neither step used outcome information: ComBat used sequence as the only batch variable (no outcome covariate) and reproducibility-based feature selection used an independent intra-/inter-observer re-segmentation set rather than the case– control labels. Radiomic discrimination was statistically equivalent to chance within a ±0.10 AUC margin (two one-sided tests). Analyses used Python (scikit-learn, neuroCombat); P < .05 was considered significant.

## Results

### Cohort

Structured reports were available for 9,956 examinations (9,808 with a parseable conus level [98.5%]; after excluding postoperative examinations the reference cohort comprised 9,655 examinations, 4,833 girls and 4,822 boys, median age 1.0 year, range 0–15). Of 1,429 children surgically treated for tethered cord in the operative registry, 571 (40.0%) linked to imaging, of whom 475 had a preoperative examination with a documented conus level.

### Reference conus position

In the reference cohort (n = 9,655), the conus medullaris terminated in the L1 region in 85.7% and the L2 region in 14.3% of children; a low-lying conus (≥L3) was present in only 0.05% (5/9,655) (Table 2). The mean level was 1.13 ± 0.33 (median L1). Conus position was marginally more cephalad with increasing age (mean 1.16 at <1 year vs 1.04 at ≥12 years; a negligible association) and nearly identical between sexes (girls 1.15, boys 1.11). All low-lying levels recovered by the improved parser among operated/postoperative cases used non-standard phrasing, so the reference-cohort low-lying rate was robust to parser recall.

**Table 1.**
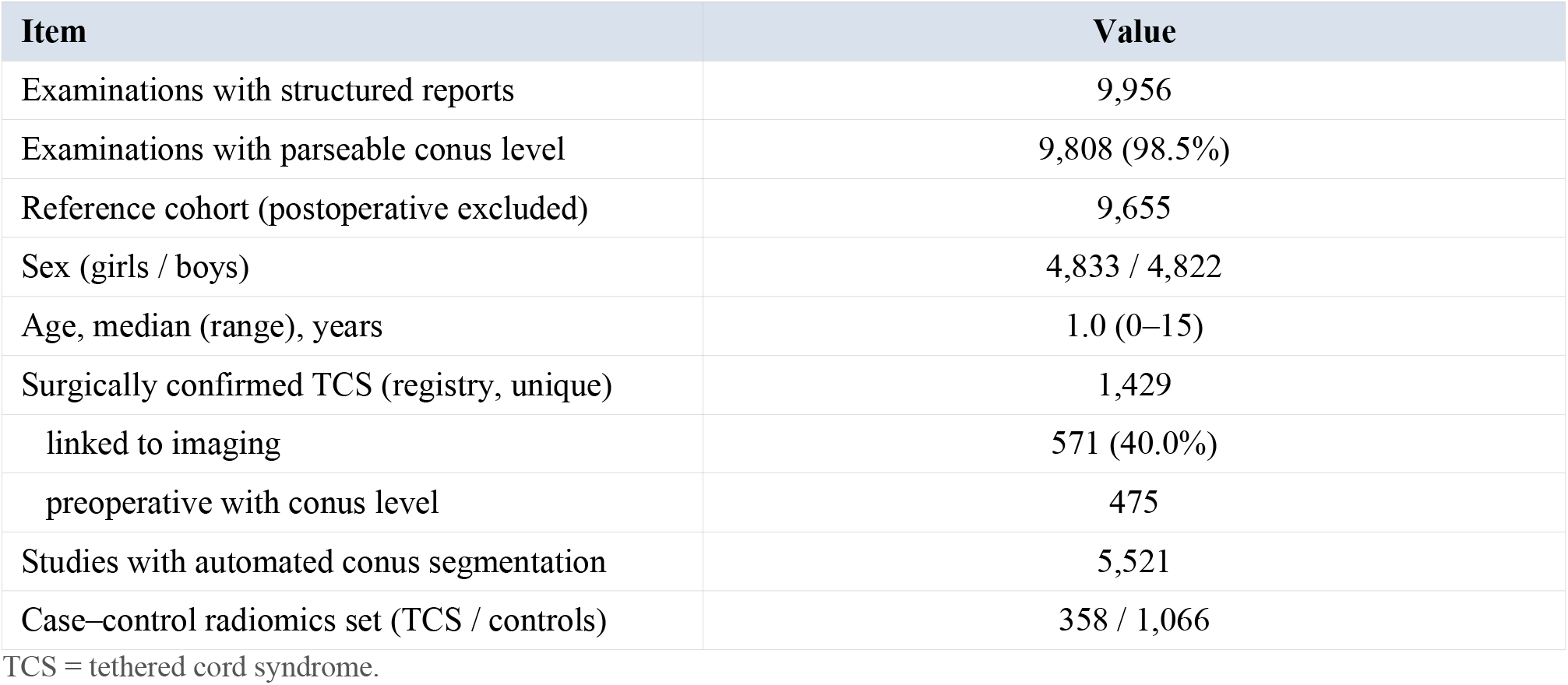
Study cohort and data sources.

| Item | Value |
| --- | --- |
| Examinations with structured reports | 9,956 |
| Examinations with parseable conus level | 9,808 (98.5%) |
| Reference cohort (postoperative excluded) | 9,655 |
| Sex (girls / boys) | 4,833 / 4,822 |
| Age, median (range), years | 1.0 (0–15) |
| Surgically confirmed TCS (registry, unique) | 1,429 |
| linked to imaging | 571 (40.0%) |
| preoperative with conus level | 475 |
| Studies with automated conus segmentation | 5,521 |
| Case–control radiomics set (TCS / controls) | 358 / 1,066 |

TCS = tethered cord syndrome.

**Table 2.** Reference distribution of conus medullaris termination level in 9,655 children.

| Group | n | Mean level ± SD* | Low-lying (≥L3) |
| --- | --- | --- | --- |
| All | 9,655 | 1.13 ± 0.33 | 0.05% |
| Age <1 y | 4,114 | 1.16 ± 0.35 | 0.10% |
| Age 1–3 y | 2,726 | 1.13 ± 0.32 | 0.00% |
| Age 3–6 y | 1,261 | 1.12 ± 0.33 | 0.08% |
| Age 6–12 y | 1,404 | 1.07 ± 0.32 | 0.00% |
| Age ≥12 y | 150 | 1.04 ± 0.29 | 0.00% |
| Girls | 4,833 | 1.15 ± 0.34 | — |
| Boys | 4,822 | 1.11 ± 0.32 | — |

\*Vertebral scale: L1 = 1, L2 = 2 (lower values = more cephalad). Distribution (reference cohort, postoperative excluded): L1 region 85.7%, L2 region 14.3%, ≥L3 0.05%.

### Conus position in surgically treated TCS

Among 475 preoperative children surgically treated for tethered cord, 99.6% had a normally positioned conus (≤L2) and only 0.4% (2 patients) were low-lying (≥L3) (Table 3). The mean conus level (1.26 ± 0.35) was only marginally lower than the reference mean. Explicit TCS-related findings were stated in the preoperative report in a minority of these patients, consistent with a radiologically occult presentation. The normal-conus finding was robust to operative subtype: the conus was normally positioned in 100% of the primary filum-type subset (n = 76) and in 99.5 –100% across all operative strata, arguing against a selection artifact driven by overt structural lesions. Matched and unmatched operative patients were comparable in sex, age, and procedure mix (standardized mean differences < 0.2), differing mainly in surgery year (more recent examinations were better represented in the imaging archive). Even under the extreme assumption that every unmatched operative patient had a low-lying conus, at least 33% of all surgically treated children would still have a normally positioned conus, versus 0.05% in the reference cohort. Explicit TCS-related findings (low conus, thickened or fatty filum) were stated in the preoperative free-text report in only a small minority of these patients (keyword-based estimate <10%), reflecting both the occult presentation and the limited sensitivity of free-text reporting for filum findings.

**Table 3.** Conus position in preoperative surgically treated TCS versus the reference cohort.

|  | Preoperative TCS | Reference cohort |
| --- | --- | --- |
| n | 475 | 9,655 |
| Mean conus level ± SD | 1.26 ± 0.35 | 1.13 ± 0.33 |
| Normally positioned (≤L2) | 99.6% | 99.9% |
| Low-lying (≥L3) | 0.4% (2) | 0.05% (5) |

TCS = tethered cord syndrome. Virtually all surgically treated TCS patients had a normally positioned conus (occult presentation).
AUC = area under the receiver operating characteristic curve. L1-penalized logistic regression, repeated stratified five-fold cross-validation, batch-only ComBat harmonization; differences by paired bootstrap.

### Automated segmentation and reproducibility

The conus segmentation model achieved a held-out Dice of 0.85 (median; mean 0.84), with a median conus-tip cranio-caudal localization error near zero but a tail of multi-slice errors reflecting the thick (median 7 mm) axial slices. Sixty-eight radiomic features were reproducible (ICC ≥ 0.75 intra- and inter-observer) and were carried forward to modeling.

### Conus texture (radiomics)

Quantitative conus texture likewise did not discriminate tethered cord from controls and was equivalent to chance; conus level was similarly uninformative. The full segmentation and radiomics pipeline, equivalence testing (nested cross-validation; AUC 0.52, equivalent within ±0.05), a positive-control detection check, and a learning-curve replication are reported in the companion methodological paper, and are not duplicated here.

**Figure 1.**
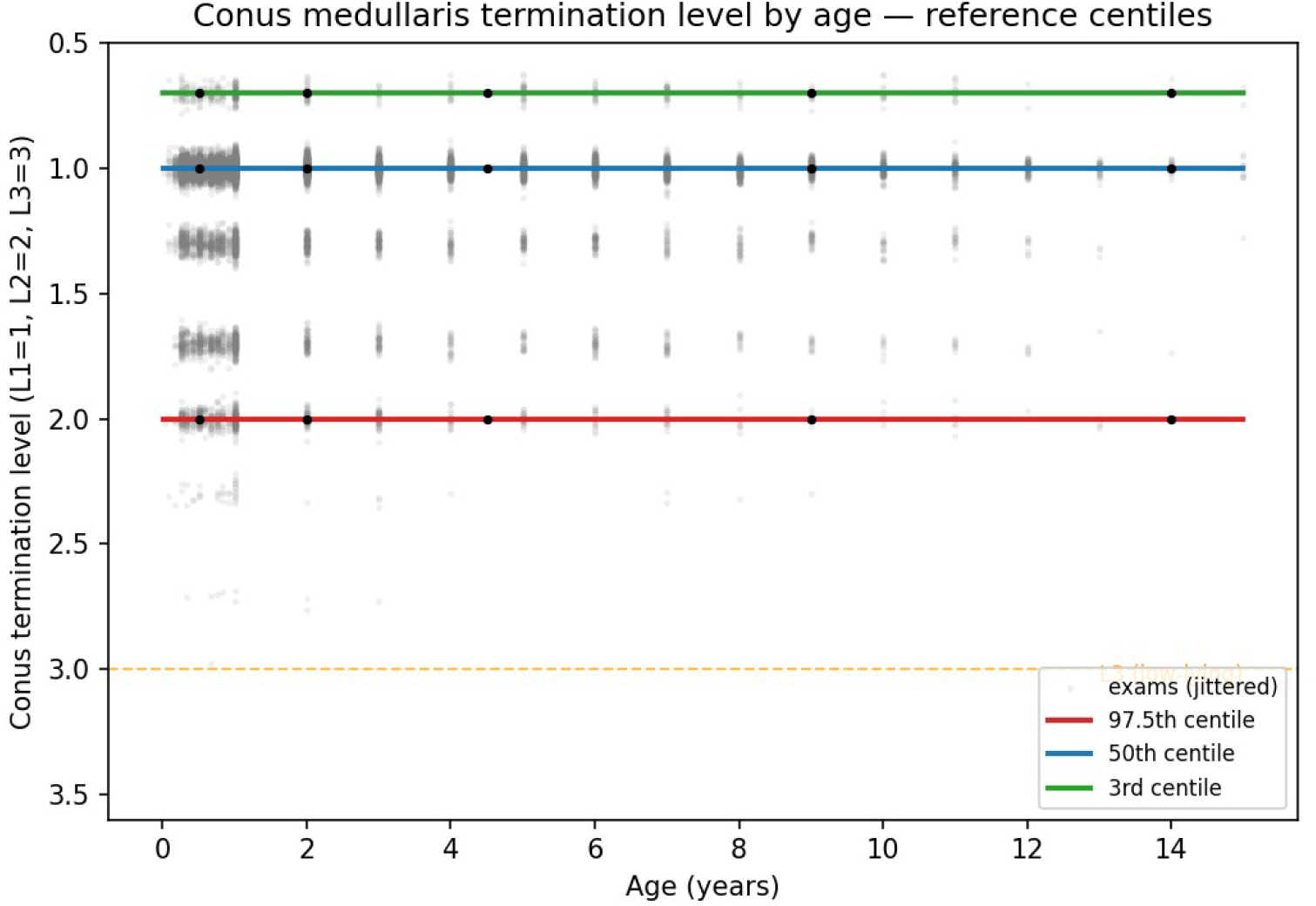
Conus medullaris termination level by age (reference cohort, postoperative examinations excluded): 3rd/50th/97.5th centiles are essentially flat across childhood, indicating an age-independent reference interval.

## Discussion

In a large pediatric cohort we found that the conus medullaris terminates at L1 or L2 in more than 99% of children and that a low-lying conus is rare (0.05%; 0.14% including operated examinations). Critically, among 475 preoperative children surgically treated for tethered cord, 99.6% had a normally positioned conus. Neither the position of the conus nor quantitative texture analysis of the conus distinguished tethered cord from normal. Together these findings indicate that, in children, the conus itself does not carry the imaging signal of tethered cord; because we did not assess the filum terminale, this does not establish that the signal lies elsewhere.

Our reference distribution (conus at L1–L2 in essentially all children, mean just below the L1 lower margin, with a slight cephalad trend across childhood) is consistent with classic anatomic descriptions but provides population-scale reference data with much narrower uncertainty than prior small series. Such reference ranges may help calibrate what should—and should not—be regarded as abnormal conus position on pediatric MRI.

The observation that virtually all surgically treated patients had a normally positioned conus supports the concept of occult tethered cord and quantifies it at scale: conus position, a historical imaging criterion, does not identify these patients. This is clinically important because it confirms, at population scale, the long-standing practice of directing diagnostic attention away from the conus and toward the clinical syndrome and the filum terminale; the filum was not quantified here and remains a hypothesis-generating candidate rather than a demonstrated alternative signal.

Consistent with the companion methodological paper, conus texture carried no reproducible signal for tethered cord (equivalence-tested null); an early small-subset signal was an acquisition artifact. We therefore do not rely on radiomics here beyond confirming that the conus—by position and by texture— does not encode the diagnosis.

Strengths of this work include the cohort scale, an automated and reproducible conus segmentation pipeline (Dice 0.85), IBSI-compliant feature extraction with explicit reproducibility filtering, harmonization without outcome leakage, and operative-registry linkage providing a surgical reference standard.

### Limitations

First, this was a single-center, retrospective study without external validation; the segmentation and analytic pipelines should be confirmed on independent data. Second, the reference distribution was derived from clinically indicated examinations, not from healthy children; the cohort is enriched for suspected spinal pathology (e.g., sacral dimple, cutaneous markers, neurogenic bladder) and skews young (median 1.0 year), so the distribution is a large-cohort clinical reference rather than a population norm. Third, conus termination level was derived from radiologist-authored reports rather than from an independent automated measurement; we attempted fully automated vertebral labeling, but routine thick-slice (3–4 mm) pediatric sagittal acquisitions limited the reliability of off-the-shelf vertebral-labeling tools, and agreement between report-stated and image-measured levels was not validated. Fourth, the operative group is a treatment cohort defined by a clinically driven surgical decision, not an independent reference standard, and was not confirmed by post-detethering outcome; registry linkage by name and date of birth may introduce occasional misclassification, and the operative list may include myelomeningocele-, lipoma-, or retethering-related procedures whose conus is abnormal by construction. Fifth, conus radiomics depended on automated segmentation (Dice 0.85), which may attenuate boundary texture; replication on manually segmented cases and an equivalence-framed power analysis are warranted. Finally, imaging reflected routine clinical protocols rather than research-optimized acquisitions. The primary-versus-secondary stratified analysis and matched-versus-unmatched comparability are reported in Results; the report-based parser was validated against blinded human report reading (weighted κ 0.946) but not against independent image-based conus measurement, which remains a limitation.

## Conclusion

In children, the conus medullaris terminates at L1–L2 in more than 99% of cases, and tethered cord treated surgically is, in essentially all cases, associated with a normally positioned conus. Neither conus position nor conus radiomics distinguishes tethered cord from normal. These large-scale, confound-controlled findings indicate that, in children, the conus itself—both its position and its texture—does not carry the imaging signal of tethered cord; they provide a large-cohort reference distribution and a validated automated conus-segmentation pipeline that motivate future study of the filum terminale and clinical phenotype. Importantly, a normally positioned conus does not exclude tethered cord and must not be used to withhold surgical evaluation.

## Data Availability

The reference distribution and analysis code are available from the corresponding author on reasonable request. Raw imaging and clinical data cannot be shared publicly owing to institutional and patient-privacy restrictions but are available from the corresponding author on reasonable request, subject to ethics approval.

## Abbreviations

TCS: tethered cord syndrome
ICC: intraclass correlation coefficient
IBSI: Image Biomarker Standardisation Initiative
AUC: area under the receiver operating characteristic curve.

## Author Contributions Statement

W.T. conceived and designed the study, performed the data analysis, and drafted the manuscript. J.C. provided imaging data and radiological interpretation. Y.D. contributed to clinical data curation and collection. Y.Y., H.H., M.Y., and J.Z. assisted with patient identification and follow-up. G.S. supervised the study and provided critical revisions. All authors reviewed and approved the final version of the manuscript.

## Ethics approval

Approved by the Ethics Committee of Women and Children’s Hospital of Ningbo University (approval no. NBFE-2026-KY-130; expedited review; approved 2 June 2026); informed consent waived (retrospective).

## Study period

Examinations performed between January 2021 and May 2026.

## Data availability

Code and feature tables available from the corresponding author / repository on reasonable request; raw imaging restricted by ethics.

Companion paper (same cohort, non-overlapping content): “Automated conus medullaris segmentation and IBSI-compliant radiomics in a large pediatric cohort.” This clinical paper reports normative conus position and the occult-TCS finding; the companion paper reports segmentation, reproducibility, radiomics, and equivalence testing.

